# The community as an active part in the implementation of interventions for the prevention and control of tuberculosis: a scoping review

**DOI:** 10.1101/2023.01.10.22283706

**Authors:** Lesly Chavez-Rimache, César Ugarte-Gil, Maria J Brunette

**Affiliations:** Instituto de Medicina Tropical Alexander von Humboldt, Universidad Peruana Cayetano Heredia, Perú; School of Medicine, Universidad Peruana Cayetano Heredia, Perú; School of Health & Rehabilitation Sciences, College of Medicine. The Ohio State University, United States of America

**Author notes:** **Corresponding author:** (CUG). **Author Contributions: Conceptualization:** Maria J Brunette. **Formal Analysis:** Lesly Chavez-Rimache, César Ugarte-Gil, Maria J Brunette. **Funding Acquisition:** Maria J Brunette. **Investigation:** Lesly Chavez-Rimache, Maria J Brunette. **Methodology:** Maria J Brunette, Lesly Chavez-Rimache. **Resources:** Lesly Chavez-Rimache, Maria J Brunette. **Supervision:** Maria J Brunette. **Visualization:** Lesly Chavez-Rimache. **Writing – Original Draft Preparation:** Lesly Chavez-Rimache, Maria J Brunette, César Ugarte-Gil. **Writing – Review & Editing:** César Ugarte-Gil, Maria J Brunette. **Competing interests:** CUG is member of the Editorial Board at PLOS Global Public Health. None of the other authors have competing interests to disclose.

**Keywords:** Community Stakeholders, Community-Based Participatory Research (CBPR), Tuberculosis

## Abstract

Interventions involving direct community stakeholders include a variety of approaches in which members take an active role in improving their health. We evaluated studies in which the community has actively participated to strengthen tuberculosis prevention and control programs. A literature search was performed in Pubmed, Scopus, ERIC, Global Index Medicus, Scielo, Cochrane Library, LILACS, Google Scholar, speciality journals, and other bibliographic references. The primary question for this review was: what is known about tuberculosis control interventions and programs in which the community has been an active part?.

Two reviewers performed the search, screening and selection of studies independently. In cases of discrepancies over the eligibility of an article, it was resolved by consensus. 130 studies were selected, of which 68.47% (n=89/130) were published after 2010. The studies were conducted in Africa (44.62%), the Americas (22.31%) and Southeast Asia (19.23%). It was found that 20% (n=26/130) of the studies evaluated the participation of the community in the detection/active search of cases, 20.77% (n=27/130) in the promotion/prevention of tuberculosis; 23.07% (n=30/130) in identifying barriers to treatment, 46.15% (n=60/130) in supervision during treatment and 3.08% (n=4/130) in social support for patient. Community participation not only strengthens the capacities of health systems for the prevention and control of tuberculosis, but also allows a better understanding of the disease from the perspective of the patient and the affected community by identifying barriers and difficulties through of the tuberculosis care cascade. Engaging key community stakeholders in co-creating solutions offers a critical pathway for local governments to eradicate TB.

## Introduction

Tuberculosis(TB) is an infectious disease that, despite being a preventable and curable disease, continues to be a problem for global public health, with a estimation of 1.6 million people died from this disease in 2021(1). In addition, tuberculosis control has become a greater challenge as it is related to other diseases such as the human immunodeficiency virus (HIV). In tuberculosis epidemiology, the social determinants of health in a community such as poverty, overcrowding, inadequate housing conditions, malnutrition, etc. they exert a clear influence on all stages of tuberculosis pathogenesis (risk of exposure, time to diagnosis, treatment, susceptibility to disease progression, and retention in care). Therefore, tuberculosis is a social disease that requires the involvement of the community to propose joint solutions with the local government(2-4).

To control TB, one of the strategies used is Directly Observed Therapy, Short Course (DOTS) (5). This strategy has been administered under the guided supervision initially by health workers, but over the years it has also included community volunteers and members of patients’ families. This has led to gradually include the community in the tuberculosis prevention and control activities. This community participation has been escalating to other levels such as the detection and active search of suspected cases, and this participation helps to generate greater linkage of patients with TB care centers and to increase the retention in care of detected cases(6).

The World Health Organization (WHO) approach called “ENGAGE-TB” mentions that community participation is fundamental to improve the scope and sustainability of tuberculosis services for the community. This approach allows the implementation of integrated community TB activities within health programs, and WHO provides technical guidance, community training, and encourages the creation and development of alliances between tuberculosis control programs and civil societies(7). Moreover, community involvement in research is encouraged, thus contributing to the strengthening of tuberculosis prevention and control programs. An example of this effort is the TB Alliance, which is an association that works with people affected by tuberculosis, who train and empower them to improve their knowledge and skills to participate in tuberculosis drug research by creating an extensive network of Community Advisory Boards (CABs). This community outreach initiative makes it possible to create links between clinical trial participants, community members and academic researchers to achieve mutual feedback based on open and consensual dialogue(8).

Community-based participatory research (CBPR) is an approach in which the community is involved in all stages of research from conception, design, execution, implementation and follow-up of research(9). CBPR allows for the creation of collaborative, equal partnerships between community members and academic researchers. Furthermore, this research approach was raised with the need to reduce communication and operational gaps that frequently imply failure for traditional research that often omits the complex cultural, social and economic interrelations and their impact on the effective implementation of interventions(9-12). CBPR can be used for all study designs, from qualitative studies to randomized clinical trials(9). To our knowledge, there are only two reviews by Arshad et al.(6) and Musa et al.(13), who evaluated the effect of community-based interventions for tuberculosis prevention and control and have reported potential benefits to include the community in TB research studies. However, it is currently unknown to what extent community involvement can generate benefits for health programs focused on tuberculosis control considering the principles of CBPR (recognize the community as a unit of identity, draw on the strengths and resources of the community, facilitate collaborative partnerships, integrate knowledge and action for the mutual benefit of all, promote a process of co-learning and community empowerment, engage a cyclical and iterative process, addressing health from a comprehensive and ecological approach, and disseminating the findings and knowledge acquired to all partners). Also, it is unknown what have been the mechanisms that have been used to strengthen communities in their participation and what has been the impact of the inclusion of the community for tuberculosis control. Based on this, our review aims to assess the characteristics of interventions in which the community has genuinely taken an active part in the development and implementation of research studies to strengthen tuberculosis prevention and control programs.

## Methods

### Study design

We carried out a scope review following the guidelines of the Preferred Reporting Items for Systematic reviews and Meta-Analyses extension for Revisión de alcances (PRISMA-ScR)(14) (Suplementary File 1) and a protocol that was carried out priori, based on the following research questions:

#### Primary question

What is known about tuberculosis control interventions and programs in which the community has been an active part in the development and implementation of the study?

#### Secondary questions

a) What strategies are being used to strengthen the participation of communities in interventions and programs for tuberculosis control? and b) What opportunities, lessons learned, and challenges impact the sustainability of community-driven interventions and programs?

The protocol is available upon request from the corresponding author. This scoping review followed the five-stage methodological framework that was developed by Arksey and O’Malley (15) and an additional sixth stage developed by Levac et al.(16), these stages are as follows: 1) identify the question research, 2) identify relevant studies, 3) select studies, 4) record data, 5) collate, summarize and report results, and 6) consult with relevant stakeholders.

### Data sources and search

To identify the studies, we systematically searched Medline (through Pubmed), Scopus, ERIC (Education Resources Information Center), Global Index Medicus, Scielo, Cochrane Library and LILACS (Latin American and Caribbean Literature in Health Sciences). A search of the gray literature was performed through Google Scholar and MedRxiv. In addition, we performed a manual search identifying the journals of internationally recognized organizations in tuberculosis (Suplementary File 2) and by reviewing the bibliographic references of the review articles and included studies. Search terms such as “Community-Based related”, “Community-based interventions”, “participatory action research”, “participatory engagement”, “Community Participation” and “tuberculosis” were used (the complete list of search terms can be found in the Suplementary File 3). The choice of terms and search strategies were developed and refined through a discussion with the research team (Suplementary File 3). For the importation of articles and the elimination of duplicates, the reference manager used was Endnote X9 (Clarivate).

### Selection criteria

This scoping review included studies that describe or analyze the role of direct involvement of key community members in the development and implementation of tuberculosis prevention and control programs. Studies such as randomized clinical trials, non-randomized trials, and observational studies were included. Letters to the editor, expert opinions, review articles, book reviews, or conference abstracts were excluded. In Figure 1 includes the processes of identification and selection of studies according to the eligibility criteria.

**Fig 1.**
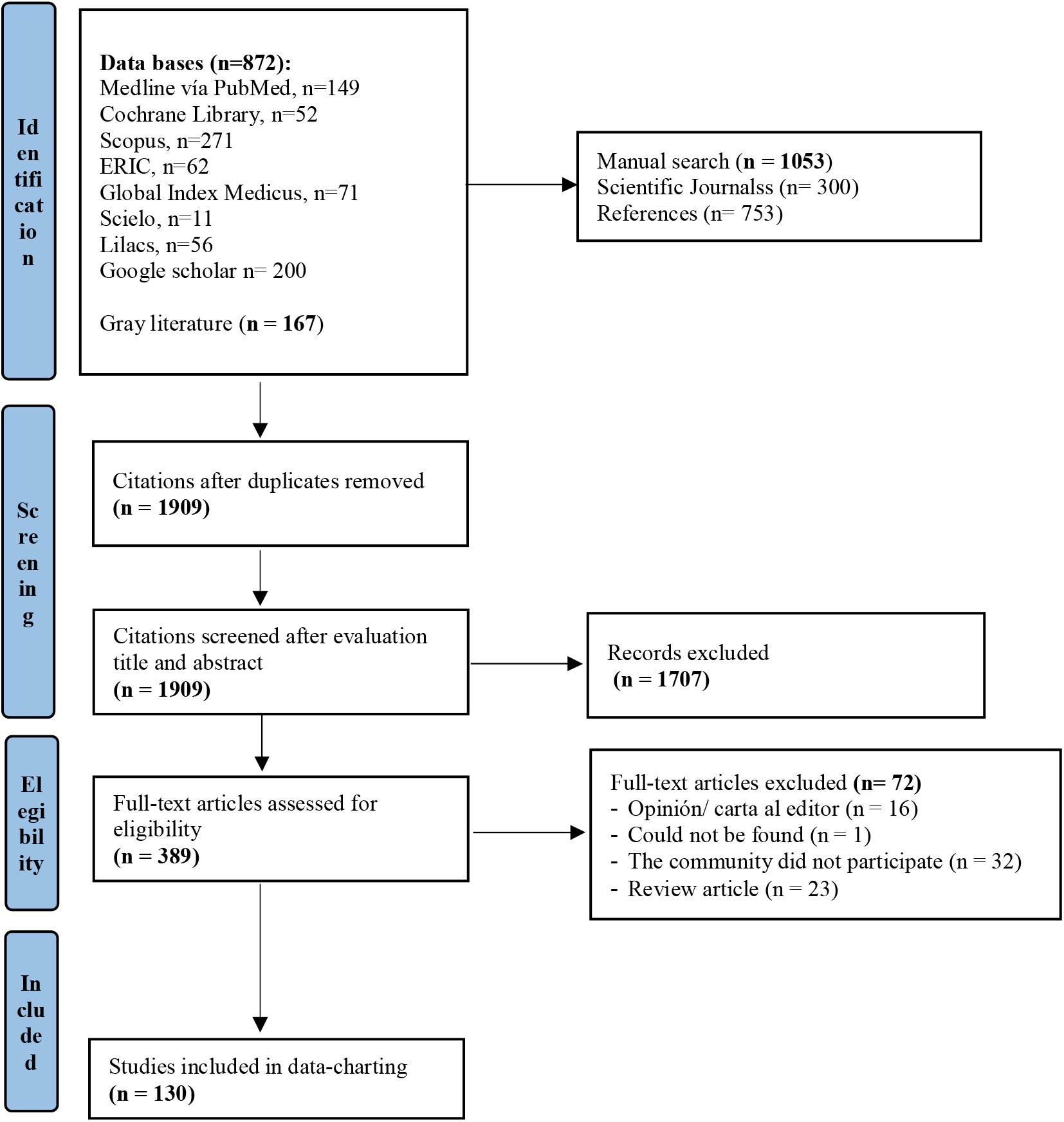
Study selection flowchart.

### Screening, collection and processing of data

One reviewer (LCR) performed initial title identification and duplicate removal using an Endnote X9 (Clarivate) reference manager. Before starting the study selection process, a calibration process was carried out between two reviewers (LCR and MJB) with 10 randomly selected studies to select the studies according to the eligibility criteria. Subsequently, in a first phase, a selection of the studies was made based on the titles and abstracts; and in a next phase, a full-text evaluation was carried out according to the eligibility criteria. All these processes were closely supervised by a researcher (MJB).

For data extraction, the “descriptive-analytic” method was used, which consists of applying a common analytical framework to all included studies and collecting standard information from each study(15). In addition, to carry out the coding of the information, a full-text evaluation of the included articles was carried out based on the research questions. The “inclusion of the community in research” and the “strengths of community participation” will be extracted from the methods section of the scientific articles and the evaluation of the “sustainability of the interventions” from the discussion section of the primary studies.

The results were presented through diagram tables that summarize the different modalities of the participation of community members in the different aspects evaluated, such as in the promotion and prevention of tuberculosis (health education, BCG vaccination and/or administration of chemoprophylaxis), detection and active search for cases, results and adherence to tuberculosis treatment (identification of barriers and difficulties for tuberculosis treatment/supervision and surveillance during tuberculosis treatment), implementation of interventions for tuberculosis control and social support and support programs for patients affected by tuberculosis. Also, the evidence was synthesized on how the community has been strengthened in its participation and finally on the challenges in the implementation of these interventions (sustainability).

## Results

### Literature search

Our scoping review initially identified 1909 titles and abstracts. Of these studies, 1707 studies were excluded based on our eligibility criteria. 389 full-text studies were evaluated, of which 130 studies finally met the pre-established inclusion criteria. The study selection flowchart is presented in Figure 1.

### Characteristics of the articles included

Of the included studies, over the years, there has been a gradual increase in the amount of research in which the community has been actively involved in tuberculosis control. The studies identified in our review were conducted in all WHO regions, mainly in the African region (44.62%, n=58/130), the Americas (22.31%, n=29/130), and in Southeast Asia (19.23%, n=25/130). In relation to the types of articles, the most frequent were the quantitative ones (46.92%, n=61/130) followed by the qualitative ones (32.31%, n=42/130) and finally the mixed methods (13.85%, n=18 /130). These characteristics are presented in Table 1.

**Table 1.**
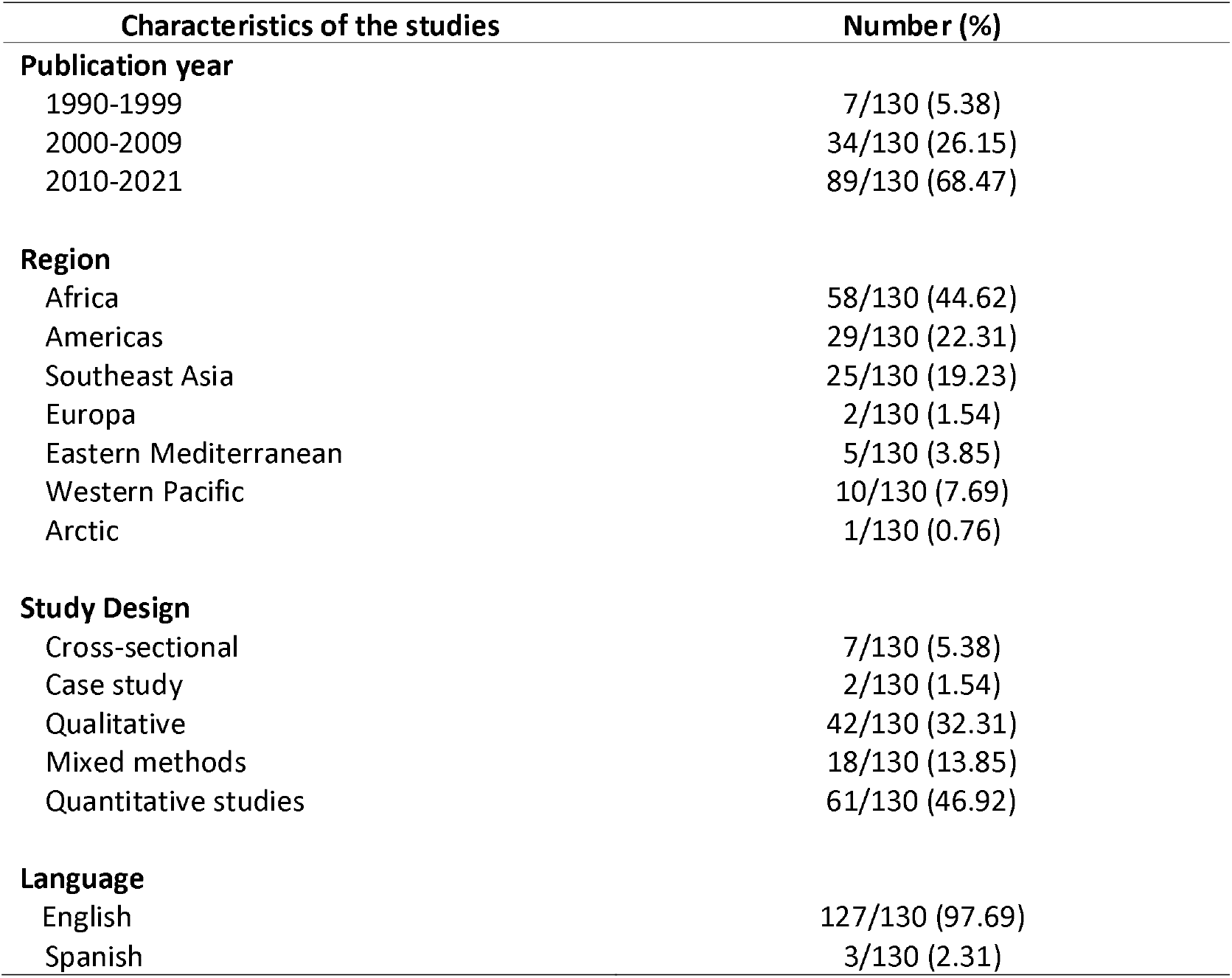
Characteristics of the studies included in the scoping review. (n=130)

### Inclusion of community participation in research

The participation of communities in research studies on tuberculosis has been evaluated and categorized considering the classification of the study by Asha et al.(17), who evaluated the degree of community participation in research on interventions in health systems in low- and middle-income countries through five different elements: (1) identification of the problems addressed; (2) identification and definition of interventions; (3) implementation of the interventions; (4) management of resources for interventions and (5) monitoring and evaluation of interventions. Only 4/130 (3.08%) studies engaged the community through these five elements. Details of the nature of participation through these five elements are presented in Table 2.

**Table 2.** Nature of community participation in the included studies.

| **Nature of Community Participation (CP)** | **Articles with ACP (n=130)**<br>**Number (%)** |
| --- | --- |
| Identification of needs, risk factors and/or definition of problems | 79/130 (60.77) |
| Identification and definition of interventions | 55/130 (42.31) |
| Implementation of interventions | 74/130 (50.92) |
| Resource management for intervention | 38/130 (29.23) |
| Monitoring, evaluation of interventions | 39/130 (30) |

ACP: Active community participation.

In the included studies, community participation in the development of research studies or as part of a program for the prevention and control of tuberculosis was identified in the following scenarios: 20.77% (n=27/130) of the studies included the community on activities related to the promotion and prevention of tuberculosis. 20.00% (n=26/130) of the studies involved the community in the detection and active search for tuberculosis cases. The 21.54% (n=28/130) of the studies in the identification of barriers and difficulties for the antituberculosis treatment. 46.15% (n=60/130) of the studies on supervision and surveillance for tuberculosis treatment and 3.08% (n=4/130) of the studies evaluated the participation of the community in social support and support for the affected patient for tuberculosis. Finally, it was identified that 12.31% (n=16/130) of the studies evaluated the participation of the community in other related aspects such as greater understanding of the disease, identification of factors related to the search and quality of care for patients. affected by tuberculosis and studies that explored the experience of community health workers in tuberculosis control (Table 3).

**Table 3.** Interventions in which the community has been an active part for the implementation of the included studies.

|  | **Articles with ACP (n=130)**<br>**Number (%)** |
| --- | --- |
| **TB prevention** | 27/130 (20.77) |
| **Detection and active search for cases** | 26/130 (20) |
| **Identification of barriers to tuberculosis treatment** | 30/130 (23.07) |
| **Supervision and surveillance for tuberculosis treatment** | 60/130 (46.15) |
| **Social support and patient support** | 4/130 (3.08) |
| **Others\*** | 16/130 (12.31) |

ACP: Active community participation.
\*Studies in which the community has participated to understand the disease, factors associated with seeking care, and experiences of community participation.

### Strengthening of community participation

6.15% (n=8/130) of the studies included in our review used the community-based participatory research approach. However, only half (n=4/130) of these studies complied with the CBPR principles. On the other hand, 37.69% (n=49/130) of the studies carried out some type of education or training for community workers to carry out their functions in tuberculosis research studies.

### Sustainability of the studies

4.62% (n=6/130) of the studies evaluated the sustainability of the interventions in which the community had participated. In addition, 2.31% (n=3/130) of the included studies evaluated the cost-effectiveness of community participation in tuberculosis care(18-20). Khan et al.(18) reported that self-administered DOTS was the most cost-effective ($164 per patient cured). However, they had a 62% cure rate as opposed to DOTS administered by community health workers (CHW) ($172 per case cured), which had a 67% cure rate. DOTS supervised by a family member ($185 per patient cured) had a cure rate of 55% and finally DOTS administered in a health center ($310 per patient cured) had a cure rate of 58%. Sinanovic et al.(19) reported that for new patients community-based DOTS was more cost-effective ($726 per successfully treated patient) than no directly observed treatment ($1201 per successfully treated patient). Similarly, for retreatment patients, community-based DOTS was more cost-effective ($1,419 per successfully treated patient) than no directly observed treatment ($2,058 per successfully treated patient). Finally, Prado et al.(20) reported that the cost per patient treated with DOTS supervised by tutors was $398 and for DOTS supervised by CHW it was $548.

## Discussion

In our scoping review, we identified 130 studies that reported genuine involvement of community members. Most of these studies were conducted in Africa (44.62%), the Americas region (22.31%), and Asia (19.23%). According to the 2021 Global Tuberculosis Report, the highest burden of tuberculosis disease occurs in the Asian and African regions (69%)(1). This could explain the reason for the higher frequency of studies found in these continents. This scenario is different for the region of the Americas, in which its global burden of tuberculosis is 2.9%, but it is the region with the second most studies that attempt to involve the community as part of their strategies to end tuberculosis. However, most of these studies are carried out in populations with a high burden of tuberculosis, such as in Latin American and Caribbean countries(21).

The results of our scoping review suggest that active community participation contributes to strengthening tuberculosis prevention and control programs. This makes visible the need to dissociate ourselves from the traditional scenario of scientific research, directed only by academic researchers, and think about addressing this disease more from a comprehensive and ecological approach that also considers the community and other social actors as keys to allow the identification of problems, and in consequence, create joint solutions that reduce the social and economic gaps that increase the burden of tuberculosis. This is similar to what was mentioned by Arshad et al.(6), who reported that community-based interventions increased the probability of tuberculosis case detection (RR: 3.1; 95% CI: 2.92 to 3.28) and success rates of treatment (RR: 1.09; 95% CI: 1.07 to 1.11). Also, they reported that the CHW, by delivering the treatment, not only increased and improved the conditions for access to care for patients affected by tuberculosis, but also improved the registration and notification systems for tuberculosis cases. Yassin et al.(22) implemented a community-based TB intervention package to bring diagnostic and treatment services closer to vulnerable communities. They reported that community participation doubled tuberculosis case notification rates and improved treatment outcomes. Similarly, other studies have reported that active case finding in countries or places with a high TB burden by CHW, community volunteers, and trained family members could strengthen notification systems and reduce gaps in access to information. health care in the community. Active case finding of smear-positive cases made an important contribution to the success on diagnosis and control of tuberculosis. This is significant because tuberculosis patients frequently present to health facilities when their disease has worsened and this is a major limitation in global efforts to control tuberculosis(23-33).

With respect to TB treatment outcomes, better results have been reported when DOTS has been delivered by CHWs, allowing for higher treatment success rates. In addition, CHWs have contributed to the supervision and follow-up for cases of drug resistance and support to patients throughout the treatment phase. Regarding the cost-effectiveness studies of DOTS provided by CHWs, Khan et al.(18) and Sinanovic et al.(19) reported that DOTS provided by CHWs was the most cost-effective for new patients. DOTS provided by the health center was found to be the least cost-effective, which is interesting as it is the WHO recommended model of practice within the current DOTS package. Khan et al.(18), reported that although self-administered treatment is the most cost-effective, it had lower cure rates compared to DOTS delivered by CHWs. On the other hand, Prado et al.(20) reported that although the DOTS performed by the tutors was the most cost effective compared to the DOTS provided by the CHWs. The DOTS carried out by the CHWs included not only providing treatment but also providing primary health education, vaccination, monitoring of risk groups and other social services in benefit of persons with TB and their families. Therefore, DOTS provided by the community demonstrates that it is an alternative that can be used by tuberculosis control programs.

In relation to the training that the CHWs received, this was very varied, but it was mainly focused on the implementation of the interventions. Only 37.7% of the studies conducted training for CHWs so that they can perform their functions. However, most of these studies only limited themselves to instructing them to perform a specific function within the research study. Reyes et al.(34) was the only included study that carried out a training program for community promoters for the prevention and control of tuberculosis in Mexico. This study empowered its promoters through educational and participatory workshops to improve understanding of the disease and about the prevention and treatment of the disease.

In our review we identified 8 studies that mentioned using the community-based participatory research (CBPR) approach. However, only 4 of these studies involved the community in all phases of the research and allowed to establish a closer link between the community and academic researchers(10, 35-37). Research with CBPR methodology is based on the principles of participatory and equitable community participation in all research processes and shared ownership of problem identification, development, and research products(12, 38-40). The fundamental principles of the CBPR approach are as follows : recognition of the community as a unique identity, builds on the strengths and resources of the community itself, promotes collaborative/equitable teamwork, facilitates mutual learning and capacity building through a process of continuous empowerment, integrates and establishes a balance between the generation of knowledge and action, a cyclical and iterative process is used, it allows evaluation by addressing health problems in the community with a comprehensive and ecological approach, it disseminates results to everyone: the academia, local governments, NGOs and community members.

These principles are adapted to the sociocultural context of the communities(9, 10). In studies with a CBPR approach, academic researchers often establish a community advisory board (or CAB), which is made up of community members who represent the voices of the community regarding their perceptions, preferences, talents, etc. and priorities. These community partners can be community health centers, public health departments, schools, prisons, and civil society organizations such as neighborhood organizations(9, 41, 42). A case of collaborative partnership with a CBPR approach is a study we conducted in the population affected by TB in Peru. We carried out a collaborative community intervention between the Universidad Peruana Cayetano Heredia, Ohio State University and the Association of Tuberculosis Patients of Comas [ASET] (Comas, district of Peru with a high rate of tuberculosis)(43). In the first phase, we carried out an empowerment program for ASET through research training for CHW in key aspects such as research methodology, research ethics, principles of the CBPR approach and on the technique and implementation of a study with the use of photovoice. In the second phase, an intervention directed by the CHW was implemented in the population affected by tuberculosis in their community through the use of the photovoice technique with the objective of evaluating the impact of the social determinants of health on tuberculosis in their community.

The inclusion of strategic research partnerships that allow the establishment of an equal relationship between the community and academic researchers requires time and financial resources(44). In low- and middle-income countries, CHWs are a key component of the health workforce to achieve the Millennium Development Goals(45). However, CHWs face many challenges such as high turnover in their functions, low motivation, inadequate supervision, lack of resources available so that they can carry out their functions, insufficient compensation or incentives, and little recognition and involvement on the part of health care providers. All of this limits their ability to contribute effectively to primary health care(46).

Among the reported functions of CHWs is providing health education (increasing awareness and knowledge about tuberculosis), detection and active search for cases, and patient follow-up(47). However, CHWs have not only been involved in the aforementioned functions, but have often solved problems that are ignored by TB programs, such as providing personalized moral comfort, nutritional support, and stigma reduction for patients affected by tuberculosis(48). Among the motivating factors for CHW are satisfaction for helping people affected by tuberculosis (feeling of prestige related to helping the neighbor), having a good relationship with health workers, respect for the community and the personal benefit they find by learning new information about tuberculosis and general health. The latter was the main motivating factor for CHWs as it empowered both them and the patients about their health and that of their community(47). In addition, CHWs can decrease the stigma of tuberculosis disease because it by drawing on their own personal experiences about the disease they can increase openness and decrease stigma among people(4, 34, 49).

The sustainability of interventions are those programs that continue to be implemented after a defined period of time while they are adapted and adapted to continue producing benefits for people(50, 51). Sustainability is an important component for the implementation and dissemination of interventions. health interventions and for the evaluation of their effects in the medium and long term. However, it is frequently poorly documented(38, 52). Lwilla et al.(53) mentioned that the DOT provided by CHWs was practical and sustainable because it allowed optimizing the time of some health workers for other duties, especially in an environment with a large increase in notification of tuberculosis cases. Dudley et al.(54) reported that at 6 years, community-based tuberculosis care was maintained despite the increasing number of patients requiring care. In addition, through this time the number of CHWs increased. However, the replicability of this model varies across community settings. In more complex urbanized settings, a barrier encountered is that health services are managed without the support of community organizations. Therefore, interventions with active community participation depend on the availability of organized community-based structures that can provide their resources and support health systems. Similarly, Wieland et al.(38) conducted a case study reporting the sustainability of a tuberculosis prevention and control program (case detection) at the adult education center of the Rochester Public Schools district.

. They report that this intervention has been sustained for 8 years due to the collaborative work of the education center, public health department, and the Rochester Healthy Community Association, whose mission is to promote health through a CBPR approach. On the other hand, Han et al.(55) conducted a descriptive study in which they mention that a community-based tuberculosis prevention program has greater potential for sustainability by training CHW to form social mobilization working groups that they can train more volunteers and allow the program to be maintained through the years. Therefore, community participation has been shown to lead to significant changes in tuberculosis control. The community’s confidence and empowerment in their own resources and capacities allows them to work differently to address complex problems such as tuberculosis.

TB programs that consider the community as an important part of their activities need to establish sustained communication (system for contacting volunteers) and frequent contact with the community organization. In addition, the CHWs must receive feedback on the progress of the program, as well as be involved in any changes or new initiatives. In addition, the empowerment of CHWs allows them to train new volunteers to carry out the activities(6).

### Strengths and limitations

This scoping review has some limitations. We made an identification of studies published in English (97.69%) and Spanish (2.31%) because the largest source of evidence is presented in those languages. We do not rule out the possibility of having studies that meet our eligibility criteria and that have been published in other languages. However, a comprehensive search was performed in recognized health databases, a gray literature search was performed, and studies were collected through the literature search and contacting authors for more information. In addition, a critical evaluation of the design of the included studies was not carried out, which limits the evaluation and characterization of the quality and certainty of the evidence that makes it possible to visualize the methodological bases of the current evidence on this topic. It would be important that this be evaluated in future systematic reviews. However, we believe that our study, by synthesizing current evidence on interventions or studies in which the community has taken an active part in their development and/or implementation, adds additional value for local and local tuberculosis prevention and control programs. global organizations consider the community as a fundamental part of the strategies to achieve their goals of eradicating tuberculosis(56).

### Future research

Based on our scoping review, we have identified that there is a paucity of information from studies that comply with the basic principles of CBPR in the population affected by tuberculosis. In addition, more studies are required to evaluate the cost effectiveness and sustainability of community participation interventions in tuberculosis control in the long term. Strategic CBPR research partnerships can mobilize and organize larger-scale community efforts to establish policies that enable social and economic policy change needed to achieve health equity(9, 57, 58).

## Conclusions

Direct community involvement in tuberculosis prevention and control in research with a community-based participatory research (CBPR) approach has not been consistently reported. However, according to our analysis, the studies found show that the active participation of the community presents a positive tendency towards the strengthening of programs for the prevention and control of tuberculosis. In addition, the participation of key members of the community not only strengthens the capacities of the health systems to generate strategies and action plans for the prevention and control of tuberculosis, but also allows a better understanding of the disease from the perspective of the patient by identifying barriers and opportunities along the cascade of care for patients affected by tuberculosis in the ‘Global South’. This scoping review also makes it possible to show that there is a need to carry out studies with the CBPR approach in the population affected by tuberculosis due to the large social component, such as the social determinants of health, that impacts this disease. This seems to indicate that studies are required that restructure the focus of a traditional investigation to one with a CBPR approach, where the population is considered as an end for the objectives of the study. The active participation of communities in co-creating solutions goes beyond the biomedical sphere and offers a critical path for regional and local governments in the fight to eradicate tuberculosis.

## Data Availability

The protocol and data is available upon request from the corresponding author.

## Acknowledgments

The authors gratefully acknowledge all the support provided by Dr. Anna Biszaha who provided training to develop this study.

## Supplementary material

S1 file. PRISMA – ScR checklist

S2 file. List of scientific journals

S3 file. Search terms and keywords

S4 file. Search strategies in each database and information repository.

