## Supplementary file 2 for "The community as an active part in the implementation of interventions for the prevention and control of tuberculosis: a scoping review"

**S2 File**. List of scientific journals

1. Bulletin of the International Union Against Tuberculosis and Lung Disease
2. Eastern Mediterranean Health Journal
3. Infectious Diseases of Poverty
4. Global Public Health
5. African Health Sciences
6. Tubercle and Lung Disease
7. American Journal of Tropical Medicine and Hygiene
8. The International Journal of Tuberculosis and Lung Disease
9. African Health Sciences
10. Social Science & Medicine
11. PLoS One
12. The Lancet Infectious Diseases
13. Revista Peruana de Medicina Experimental y Salud Pública
14. Global Health: Science and Practice
15. Community Development Journal
16. Revista de Saúde Pública
17. Journal of Epidemiology and Community Health
18. African Journal of Primary Health Care & Family Medicine
19. Nigerian Journal of Medicine
20. Eastern Mediterranean Health Journal
21. Canadian Journal of Public Health
22. International Journal of Public Health Research
23. International Journal of Environmental Research and Public Health
24. Journal of Health Organization and Management
25. BMC Public Health
26. Public Health
27. Journal of Health Communication
