## Supplementary file 3 for "The community as an active part in the implementation of interventions for the prevention and control of tuberculosis: a scoping review"

**S3 file.** Search terms and keywords

Search terms

Community-Based related

"Community-based interventions"

"participatory action Research"

"participatory research"

"participatory engagement"

"community research"

"action research"

"Community-Based Participatory Research (CBPR)"

"Participation, Community"

"Community Participation" (Mesh)

"Community Involvement"

"Community Involvements"

"Involvement, Community"

"Involvements, Community"

"Consumer Participation"

"Participation, Consumer"

"Consumer Involvement"

"Consumer Involvements"

"Involvement, Consumer"

"Public Participation"

"Participation, Public"

"Community Action"

"Action, Community"

"Actions, Community"

"Community Actions"

Tuberculosis

"Tuberculosis"

"Tuberculoses"

"Kochs Disease"

"Koch's Disease"

"Koch Disease"

((("Tuberculosis/prevention and control"[Mesh]) OR "Tuberculosis"[Mesh]) OR "Tuberculosis/transmission"[Mesh]) OR (Tuberculosis/epidemiology"[Mesh] OR "Tuberculosis/organization and administration"[Mesh])

"Mycobacterium tuberculosis Infection"

"Infection, Mycobacterium tuberculosis"

"Infections, Mycobacterium tuberculosis"

"Mycobacterium tuberculosis Infections"

"Tuberculosis/Diagnostics"

"Tuberculosis/Treatment adherence"

"Tuberculosis/Control"

"Tuberculosis/Prevention"
