## Supplementary file 4 for "The community as an active part in the implementation of interventions for the prevention and control of tuberculosis: a scoping review"

**S4 File.** Search strategies in each database and information repository.

| Databases | Search strategies |
| --- | --- |
| Medline (Pubmed) | ("Community-based interventions"[TIAB] OR "participatory action Research"[TIAB] OR "participatory research" [TIAB] OR "participatory engagement" [TIAB] OR "community research" [TIAB] OR "action research" [TIAB] OR "Community-Based Participatory Research (CBPR)" [TIAB] OR "Participation, Community" [TIAB] OR "Community Participation" [Mesh] OR "Community Involvement"[TIAB] OR "Community Involvements"[TIAB] OR "Involvement, Community"[TIAB] OR "Consumer Participation"[TIAB] OR "Participation, Consumer"[TIAB] OR "Consumer Involvement" [TIAB] OR "Involvement, Consumer"[TIAB] OR "Public Participation"[TIAB] OR "Participation, Public"[TIAB] OR "Community Action"[TIAB] OR "Action, Community"[TIAB] OR "Actions, Community"[TIAB] OR "Community Actions"[TIAB]) AND ("Tuberculosis"[TIAB] OR "Tuberculoses"[TIAB] OR "Koch's Disease"[TIAB] "Tuberculosis"[Mesh] OR "Mycobacterium tuberculosis Infection"[TIAB] OR "Infection, Mycobacterium tuberculosis"[TIAB] OR "Infections, Mycobacterium tuberculosis"[TIAB] OR "Mycobacterium tuberculosis Infections"[TIAB] OR "Tuberculosis/Diagnostics"[TIAB] OR "Tuberculosis/Treatment adherence"[TIAB] OR "Tuberculosis/Control"[TIAB] OR "Tuberculosis/Prevention"[TIAB] OR ((("Tuberculosis/prevention and control"[Mesh]) OR "Tuberculosis"[Mesh]) OR "Tuberculosis/transmission"[Mesh]) OR ("Tuberculosis/epidemiology"[Mesh] OR "Tuberculosis/organization and administration"[Mesh])) |
| Scopus | TITLE-ABS-KEY ("Community-based interventions" OR "participatory action Research" OR "participatory research" OR "participatory engagement" OR "community research" OR "action research" OR "Community-Based Participatory Research (CBPR)" OR "Participation, Community" OR "Community Participation" OR "Community Involvement" OR "Community Involvements" OR "Involvement, Community" OR "Involvements, Community" OR "Consumer Participation" OR "Participation, Consumer" OR "Consumer Involvement" OR "Consumer Involvements" OR "Involvement, Consumer" OR "Public Participation" OR "Participation, Public" OR "Participation, Public" OR "Action, Community" OR "Actions, Community" OR "Community Actions" OR "Community-Based Health" ) AND TITLE-ABS-KEY ( "Tuberculosis" OR "Tuberculoses" OR "Kochs Disease" OR "Koch's Disease" OR "Mycobacterium tuberculosis Infection" OR "Infection, Mycobacterium tuberculosis" OR "Infections, Mycobacterium tuberculosis" OR "Mycobacterium tuberculosis Infections" OR "Tuberculosis/Diagnostics" OR "Tuberculosis/Treatment adherence" OR "Tuberculosis/Control" OR "Tuberculosis/Prevention") |
| ERIC | (Community-Based Participatory Research OR Community Participation) AND (Tuberculosis) |
| LILACS | ("Community-Based related" OR "Community-based interventions" OR "participatory action Research" OR "participatory research" OR "participatory engagement" OR "community research" OR "action research" OR "Community-Based Participatory Research (CBPR)" OR "Participation, Community" OR "Community Participation" OR "Community Involvement" OR "Community Involvements" OR "Involvement, Community" OR "Involvements, Community" OR "Consumer Participation" OR "Participation, Consumer" OR "Consumer Involvement" OR "Consumer Involvements" OR "Involvement, Consumer" OR "Public Participation" OR "Participation, Public" OR "Participation, Public" OR "Action, Community" OR "Actions, Community" OR "Community Actions" OR "Community-Based Health") AND ("Tuberculosis" OR "Tuberculoses" OR "Kochs Disease" OR "Koch's Disease" OR "Mycobacterium tuberculosis Infection" OR "Infection, Mycobacterium tuberculosis" OR "Infections, Mycobacterium tuberculosis" OR "Mycobacterium tuberculosis Infections" OR "Tuberculosis/Diagnostics" OR "Tuberculosis/Treatment adherence" OR "Tuberculosis/Control" OR "Tuberculosis/Prevention") |
| GLOBAL INDEX MEDICUS | ("Community-Based related" OR "Community-based interventions" OR "participatory action Research" OR "participatory research" OR "participatory engagement" OR "community research" OR "action research" OR "Community-Based Participatory Research (CBPR)" OR "Participation, Community" OR "Community Participation" OR "Community Involvement" OR "Community Involvements" OR "Involvement, Community" OR "Involvements, Community" OR "Consumer Participation" OR "Participation, Consumer" OR "Consumer Involvement" OR "Consumer Involvements" OR "Involvement, Consumer" OR "Public Participation" OR "Participation, Public" OR "Participation, Public" OR "Action, Community" OR "Actions, Community" OR "Community Actions" OR "Community-Based Health") AND ("Tuberculosis" OR "Tuberculoses" OR "Kochs Disease" OR "Koch's Disease" OR "Mycobacterium tuberculosis Infection" OR "Infection, Mycobacterium tuberculosis" OR "Infections, Mycobacterium tuberculosis" OR "Mycobacterium tuberculosis Infections" OR "Tuberculosis/Diagnostics" OR "Tuberculosis/Treatment adherence" OR "Tuberculosis/Control" OR "Tuberculosis/Prevention") |
| Scielo | ("Community-based interventions" OR "participatory action Research" OR "participatory research" OR "participatory engagement" OR "community research" OR "action research" OR "Community-Based Participatory Research (CBPR)" OR "Participation, Community" OR "Community Participation" OR "Community Involvement" OR "Community Involvements" OR "Involvement, Community" OR "Involvements, Community" OR "Consumer Participation" OR "Participation, Consumer" OR "Consumer Involvement" OR "Consumer Involvements" OR "Involvement, Consumer" OR "Public Participation" OR "Participation, Public" OR "Participation, Public" OR "Action, Community" OR "Actions, Community" OR "Community Actions" OR "Community-Based Health") AND ("Tuberculosis" OR "Tuberculoses" OR "Kochs Disease" OR "Koch's Disease" OR "Mycobacterium tuberculosis Infection" OR "Infection, Mycobacterium tuberculosis" OR "Infections, Mycobacterium tuberculosis" OR "Mycobacterium tuberculosis Infections" OR "Tuberculosis/Diagnostics" OR "Tuberculosis/Treatment adherence" OR "Tuberculosis/Control" OR "Tuberculosis/Prevention") |
| Cochrane Library | ID Search Hits  #1 MeSH descriptor: [Community Participation] explode all trees 1713  #2 (Community-based interventions):ti,ab,kw 2693  #3 (participatory action Research):ti,ab,kw 163  #4 (participatory research):ti,ab,kw 984  #5 (participatory engagement):ti,ab,kw 177  #6 (community research):ti,ab,kw 12104  #7 (action research):ti,ab,kw 5471  #8 (Community-Based Participatory Research (CBPR)):ti,ab,kw 151  #9 (Participation, Community):ti,ab,kw 4031  #10 (Community Involvement):ti,ab,kw 1080  #11 (Consumer Participation):ti,ab,kw 306  #12 (Consumer Involvement):ti,ab,kw 110  #13 (Consumer Involvements):ti,ab,kw 1  #14 (Public Participation):ti,ab,kw 1558  #15 (Community Action):ti,ab,kw 1433  #16 (Actions, Community):ti,ab,kw 374  #17 MeSH descriptor: [Tuberculosis] explode all trees 2058  #18 (Tuberculoses):ti,ab,kw 17  #19 (Kochs Disease):ti,ab,kw 1  #20 (Koch's Disease):ti,ab,kw 48  #21 (Koch Disease):ti,ab,kw 48  #22 (Mycobacterium tuberculosis Infection):ti,ab,kw 382  #23 (Infection, Mycobacterium tuberculosis):ti,ab,kw 382  #24 (Infections, Mycobacterium tuberculosis):ti,ab,kw 213  #25 (#1 OR #2 OR #3 OR #4 OR #5 OR #6 OR #7 OR #8 OR #9 OR #10 OR #11 OR #12 OR #13 OR #14 OR #15 OR #16) AND (#17 OR #18 OR #19 OR #20 OR #21 OR #22 OR #23 OR #24) |
| Google Scholar | ("Community-based" OR "participatory action Research" OR "participatory research" OR "Community engagement" OR "Community-Based Participatory Research" OR "Community Participation" OR "Capacity Building") AND ("Tuberculosis") |
